# Uptake, reidentification and regret in pediatric gender care

**DOI:** 10.64898/2026.09.17.26363359

**Authors:** Michelle A. Tollit, Will Conabere, Katherine Gray, Chelsea Sibley, Izza Ayub, Carmen C. Pace, Anna Traill, Tessa Russo-Batterham, Blake S. Cavve, Tanya Ravipati, Cate Rayner, Ken C. Pang

## Abstract

**Background:** Gender-affirming medical care (GAMC) for children and adolescents in Australia has become increasingly politicized, despite care being provided in accordance with latest evidence-based international guidelines. Critics of GAMC claim that current assessment and deliberation processes are insufficient, and that too many young people are accessing medical interventions, no longer identifying as trans, and expressing treatment regret. These claims have led to widespread bans on the provision of pediatric GAMC, despite growing evidence regarding harms caused by such bans.

**Aim:** This study sought to describe the care journeys of the young people attending Australia’s largest pediatric gender service, with a focus on reporting the rates at which young people commenced medical interventions, re-identified with their birth-assigned sex, and expressed regret about their treatment.

**Methods:** The Royal Children’s Hospital Gender Service (RCHGS) is a statewide service that provides individualised multidisciplinary care to trans and gender diverse young people in Victoria, Australia. An initial extended triage appointment explores the needs and goals of the young person and their family. Some proceed to receive individualised multidisciplinary care (MDC) at RCHGS which can include psychosocial assessment and management, family support, and medical interventions including pubertal suppression (PS) and gender-affirming hormonal therapy (GAHT). We undertook a retrospective file audit of all patients who had attended at least one RCHGS appointment and been discharged between 1/1/2017- 31/12/2024. For each person, key data were extracted from their electronic medical record and recorded in REDCap.

**Results:** 1,391 patients were discharged over the 8-year study period and many different care trajectories were observed. 814 (58.5%) did not access ongoing MDC. Of the remaining 577, 353 (25.4% of total cohort) started a medical intervention ((PS: n=64/1391 (4.6%); estradiol: n=97/1391 (7.0%); testosterone: n=244/1391 (17.5%)). For those who started these medical interventions, the median duration and number of sessions from first appointment until commencement of medications was 530 (IQR: 371–693) days and 9 (IQR: 7–13) sessions. 346 of these patients (98.0%) had data on gender identity at discharge and subsequent care pathways. 7 (2.0%) re-identified with their birth-registered sex and stopped treatment: 4 did so after commencing PS (4/61=6.6% of those starting PS), 2 did so after commencing testosterone, and 1 did so after commencing estradiol (3/341=0.9% of those starting GAHT). Just one of these patients, who had been on estradiol for 1 month, expressed treatment regret given concerns about initial breast budding and future fertility.

**Conclusion:** This study highlights varied trajectories of those referred to pediatric gender services, directly challenges claims used to justify restrictions on pediatric GAMC and underscores the importance of a person-centred approach to care.

## Introduction

As in other countries, gender-affirming medical care (GAMC) for children and adolescents in Australia has become increasingly politicized, despite care being provided in accordance with latest evidence-based international guidelines.^1^ Notably, flawed and outdated historical data have been used by critics to justify the expansion of restrictions on pediatric GAMC,^2^ despite growing evidence regarding harms caused by such bans.^3^ While trans young people have highlighted the impact of lengthy assessment processes,^4^ critics of GAMC claim that current assessment and deliberation processes are insufficient,^5^ and that too many young people are accessing medical interventions, no longer identifying as trans, and expressing treatment regret.^6^ In this study, we aim to describe the care journeys of young people attending Australia’s largest pediatric gender service, with a focus on the rates at which young people commenced medical interventions, re-identified with their birth-assigned sex, and expressed regret about their treatment.

## Methods

The Royal Children’s Hospital Gender Service (RCHGS) provides individualised multidisciplinary care to transgender or gender diverse young people throughout Victoria, Australia, up to age 18 years. An initial extended triage appointment provides opportunity to explore the needs and goals of the young person and their family. For some, further care at RCHGS is not accessed and they are discharged, often to community-based supports. Others proceed to receive individualised multidisciplinary care (MDC) at RCHGS – usually after being put onto a secondary waitlist – which can include psychosocial assessment and management, family support, and medical interventions including pubertal suppression (PS) with gonadotropin releasing hormone agonists and gender-affirming hormonal therapy (GAHT) with estradiol and testosterone.

We undertook a retrospective file audit of all patients who had attended at least one RCHGS appointment and been discharged between 1/1/2017-31/12/2024 (ethics approval #36323). Key data were extracted from each patient’s electronic medical record and recorded in REDCap (see Supplementary Materials for protocol). Descriptive statistics were calculated.

## Results

1,391 patients (Table 1) were discharged over the 8-year study period and many different care trajectories were observed (Figure 1). 814 (58.5%) did not access ongoing MDC. Of the remaining 577, 353 (25.4% of total cohort) started a medical intervention ((PS: n=64/1391 (4.6%); estradiol: n=97/1391 (7.0%); testosterone: n=244/1391 (17.5%)). For those who started these medical interventions, the median duration and number of sessions from first appointment until commencement of medications was 530 (IQR:371–693) days and 9 (IQR:7–13) sessions (Table 1). 346 of these patients (98.0%) had data on gender identity at discharge and subsequent care pathways. 7 (2.0%) re-identified with their birth-registered sex and stopped treatment: 4 did so after commencing PS (4/61=6.6% of those starting PS), 2 did so after commencing testosterone, and 1 did so after commencing estradiol (3/341=0.9% of those starting GAHT). Just one of these patients, who had been on estradiol for 1 month, expressed treatment regret given concerns about initial breast budding and future fertility.

**Table 1.** Characteristics of patients discharged over the 8-year study period (n=1391)

|  | Frequency (%)<br>(unless otherwise specified) |
| --- | --- |
| **Age at first appointment, n=1391 (100.0%)** |  |
| Median (IQR) | 15.5 (13.9–16.5) |
| **Age at discharge from service, n=1391 (100.0%)** |  |
| Median (IQR) | 17.3 (15.7–18.3) |
| **Gender identity (at first appointment), n=1391 (100.0%)** |  |
| Trans masculine | 644 (46.3%) |
| Trans feminine | 286 (20.6%) |
| Non-binary | 315 (22.6%) |
| Cis male | 25 (1.8%) |
| Cis female | 13 (0.9%) |
| Young person unsure | 101 (7.3%) |
| Prefer not to say | 1 (0.1%) |
| Unclear to auditor | 6 (0.4%) |
| **Sex assigned at birth, n=1391 (100.0%)** |  |
| Male | 414 (29.8%) |
| Female | 977 (70.2%) |
| **IRSAD Quintile<sup>a</sup>, n=1368 (98.3%)** |  |
| Quintile 1 | 143 (10.5%) |
| Quintile 2 | 185 (13.5%) |
| Quintile 3 | 253 (18.5%) |
| Quintile 4 | 354 (25.9%) |
| Quintile 5 | 433 (31.7%) |
| **Rurality – Modified Monash Model<sup>b</sup>, n=1368 (98.3%)** |  |
| Metropolitan | 939 (68.6%) |
| Regional centre | 162 (11.8%) |
| Large rural town | 56 (4.1%) |
| Medium rural town | 37 (2.7%) |
| Small rural town | 174 (12.7%) |
| **Aboriginal and/or Torres Strait Islander identity, n=1288 (92.6%)** |  |
| Aboriginal and/or Torres Strait Islander | 35 (2.7%) |
| Non-Aboriginal or Torres Strait Islander | 1253 (97.3%) |
| **Language spoken at home, n=1369 (98.4%)** |  |
| English | 1320 (96.4%) |
| Language other than English | 49 (3.6%) |
| **Country of birth/migration, n=1370 (98.5%)** |  |
| Born in Australia | 1282 (93.6%) |
| Born overseas | 88 (6.4%) |
| **Duration of care at RCHGS, n=1391 (100.0%)** |  |
| Duration (in days) from first appointment to discharge – Median (IQR) | 580 (138–953) |
| **Duration, in days, (median [IQR]) from first appointment until commencement of GAMC, n=353 (100%)<sup>c</sup>** |  |
| First appointment to commencement of GAMC (n=353) | 530 (371–693) |
| First appointment to commencement of PB (n=64) | 207 (109–449) |
| First appointment to commencement of estradiol (n=97) | 637 (427–980) |
| First appointment to commencement of testosterone (n=244) | 574 (437–754) |
| **No. sessions (median [IQR]) from first appointment until commencement of GAMC, n=353 (100%)<sup>d</sup>** |  |
| First appointment to commencement of GAMC (n=353) | 9 (7–13) |
| First appointment to commencement of PB (n=64) | 6 (5–10) |
| First appointment to commencement of estradiol (n=97) | 14 (10–19) |
| First appointment to commencement of testosterone (n=244) | 10 (8–13) |

**Figure 1.**
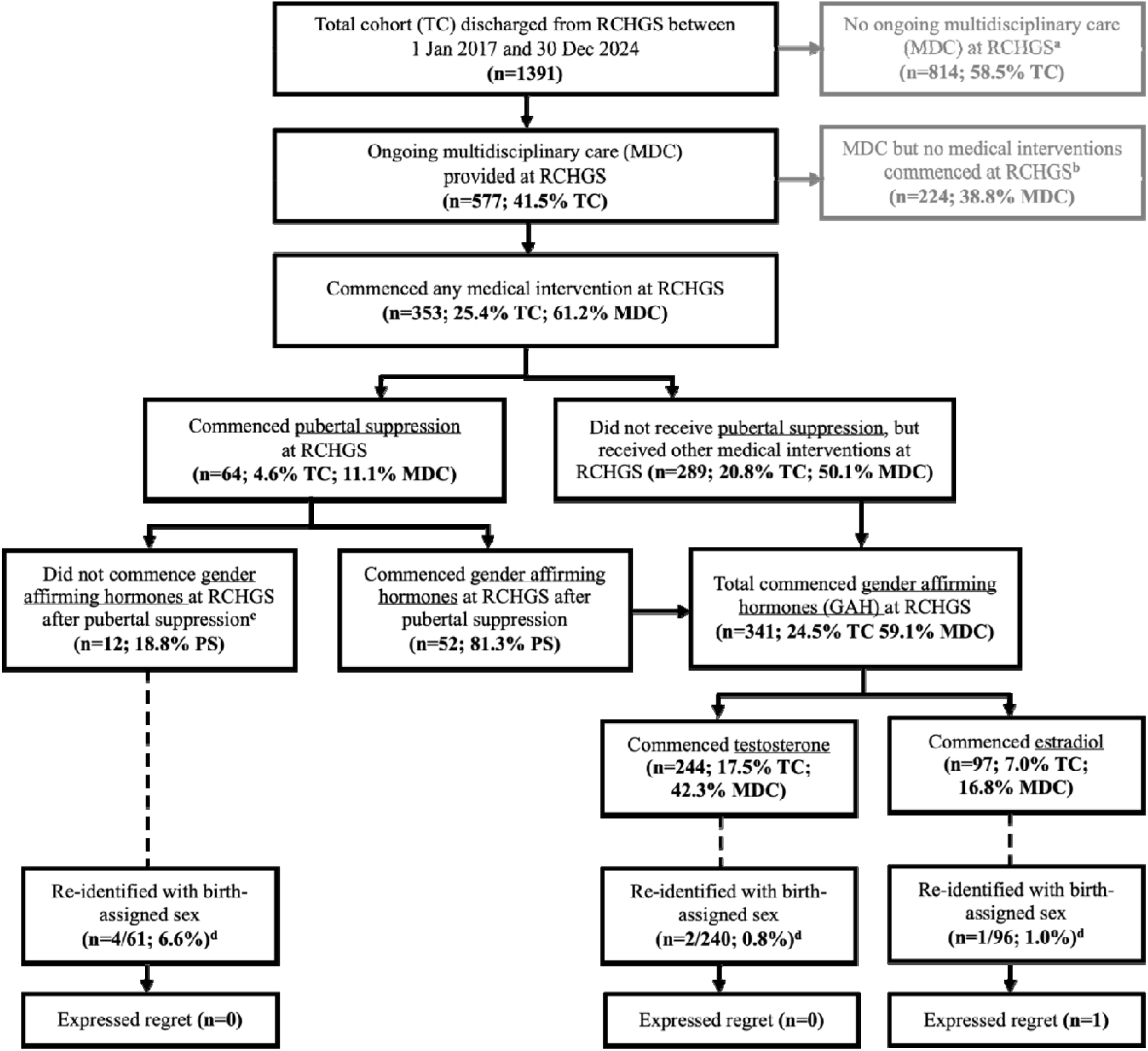
Rates of medical intervention uptake, re-identification and regret within a cohort of young people attending a pediatric gender service. This flowchart summarises clinical pathways and outcomes of 1,391 young people discharged from the RCHGS during the 8-year period from 1/1/2017 until 31/12/2024. The numbers of young people who started pubertal suppression (PS), testosterone and estradiol through RCHGS are shown, and rate of uptake is presented as a percentage of the total number of discharges (i.e, the total cohort (TC)) and as a percentage of those who commenced medical interventions (MI) at RCHGS. All proportions were performed on complete cases. ^a^ 814 young people were discharged with no ongoing multidisciplinary care (MDC) at RCHGS. This includes those patients discharged after an initial triage appointment, those who did not attend any additional appointments (including those lost to follow-up and those who declined further appointments), and those who were managed through the Under 8 pathway without progression to MDC. ^b^ Participants where it was unclear whether medical interventions were initiated at RCHGS (n=1) or if they initiated these outside of RCHGS (n=13) were excluded from the MI sub-cohort analysis. ^c^ For these 12 patients, 4 re-identified with birth-registered sex, 3 moved interstate and continued GAMC, 3 shifted in gender identity (non-binary, n=2; unsure, n=1) and no longer requested a medical pathway, and 2 were unclear/lost to follow up. ^d^ Rates of re-identification are reported as a percentage of those who commenced each type of medical intervention, with denominators restricted to those with complete data on gender identity at discharge and subsequent care pathway. Participants with a missing or unclear gender identity at discharge (n=4) or unclear subsequent care pathway (n=3) were excluded from the re-identification rate denominators but are still included in the medical intervention totals.

## Discussion

Our findings highlight differing care trajectories following attendance at a pediatric gender service. Most young people did not access ongoing MDC, instead undergoing a single consultation providing education and linkages to community supports. This is consistent with recent reductions in RCHGS referrals, as more community-based support options have become available.^7^

A minority of young people commenced medical interventions, often after numerous appointments over years, reflecting a considered approach. Timeframes for accessing GAMC varied, illustrating the diversity of care pathways. For example, young children are not eligible for medical interventions until many years after service entry, while others can experience long wait times for MDC. It is important to acknowledge that the latter – in conjunction with high appointment numbers – can be distressing for trans young people and their families,^4^ and balancing timely access to care with available resources and patient safety is a key challenge for pediatric gender services.

In keeping with other recent data,^8^ we found young people who accessed GAMC rarely reidentified with their birth-registered sex. Importantly, re-identification per se is not undesirable, especially given PS aims to provide more time for young people to explore their gender.

Of those who re-identified with birth-registered sex, we identified only a single case of treatment regret, representing a considerably lower rate than other areas of pediatrics.^9^ Notably, regret centred around initial breast budding, which would be expected to resolve upon estradiol cessation, and concerns about long-term fertility, which recent data suggest are unwarranted.^10^

Our study’s limitations require acknowledgement. Findings are from a single service, limiting generalisability. They were also restricted to information in the medical record, and didn’t feature information after discharge, leaving the possibility that outcomes may have subsequently changed.

In conclusion, our study highlights varied trajectories of those referred to pediatric gender services, directly challenges claims used to justify restrictions on pediatric GAMC and underscores the importance of a person-centred approach to care. Looking ahead, future research should focus on supporting evidence-based models of GAMC that are equitable and timely.

## Data Availability

Data requests, accompanied by a methodologically sound proposal can be submitted to the corresponding author.

## Acknowledgements

We wish to acknowledge Dr Tram Nguyen, Dr Kevin Conlon, Dr Lou Kerley and Dr Pip Buckingham for their helpful assistance with this project. No compensation was received for their role in this project.

## Data Sharing Statement

Data requests, accompanied by a methodologically sound proposal outlining their scientific use of these data, can be submitted to the corresponding author, KP, for consideration. If the request is approved, data will only be shared once relevant institutional ethics approvals have been obtained.

