## Supplement for "Uptake, reidentification and regret in pediatric gender care"

**SUPPLEMENTAL MATERIALS**

**eMethods**

**eCodebook**

**eTable 1 – Data extracted related to primary outcomes**

**eTable 2 – Data extracted related to other outcomes**

**eTable 3 – Supplementary data extracted**

**eReferences**

**eMethods**

**1.0 Objective**

The goal of this audit was to document the treatment journey of those who attend the Royal Children’s Hospital Gender Service (RCHGS), including key data relating to uptake and cessation of medical intervention/s at RCHGS, re-identification with birth-assigned sex, and regret regarding medical intervention/s in children and adolescents who had been discharged from the RCHGS.

**2.0 Study type**

Retrospective medical record audit in a cohort of children and adolescents who had been discharged from RCHGS.

**3.0 Setting**

The Royal Children’s Hospital Gender Service (RCHGS) was formally established in 2012/2013^[1]^ and is currently Australia’s largest pediatric gender service. This service delivers individualised, multi-disciplinary, evidence-based gender-affirming care.

Individuals aged 8 years and older at service entry are initially seen in a Single Session Nurse led assessment clinic (SSNac). This is a 90-minute appointment focused on health education, the provision of information on community support options, and triage. Following a secondary waiting period, patients may attend a multidisciplinary clinic (MDC) with a psychologist/psychiatrist, a pediatrician/endocrinologist and other allied health if needed (such as a speech pathologist or social worker). The MDC consists of a series of appointments, covering medical, developmental and psychosocial background, and goals of care. Throughout the MDC, individual and family support is provided and if relevant, gender affirming medications are prescribed and monitored.

Individuals under 8 years of age at service entry attend appointments with a mental health clinician only; when they approach puberty, they may also be seen by a pediatrician/endocrinologist alongside a mental health professional for assessment, support, and if indicated, gender affirming hormones, consistent with the MDC.

Young people can remain with the service until they are 18 years old after which they are discharged. Other reasons for being discharged (at any point throughout their journey with RCHGS) include not requiring medical care and/or if they indicate they do not identify as trans or gender diverse, or if they seek care elsewhere (in which case their care may be transferred to other services, including General Practitioners (GPs), external psychologists, or adult services). If patients do not attend multiple appointments, and are unable to be contacted, they may also be discharged from the service.

**4.0 Participants (analytic sample)**

**Inclusion criteria**

All patients discharged between 1 January 2017 and 31 December 2024 who had attended at least one gender-related or RCHGS appointment through the Royal Children’s Hospital. For the purpose of this audit, a patient was considered ‘discharged’ if they were recorded as such in the RCHGS administrative records.

**Exclusion criteria**

Patients whose primary health service for gender-related care was not the RCHGS. This includes a small number of patients receiving care in other regions, who accessed the RCHGS for a specific purpose, such as bone density scanning, but ongoing health care occurred elsewhere, not at the RCH.

**5.0 Outcomes**

**Primary outcomes (subject of current paper)**

Commencement of medical intervention/s at RCHGS

Cessation of medical intervention/s

Re-identification with birth-assigned sex

Regret regarding medical intervention/s

**6.0 Procedure: Data Collection - Extraction / Exports**

A multipronged approach was used to extract data as described below.

Discharged patients were initially identified from the RCHGS’s administrative records. This included patients who no longer required ongoing care, reached the age threshold, or who were lost to follow up.

**Medical record audit – Manual data extraction**

A data collection form was developed along with a detailed codebook for each variable to ensure consistency between auditors. For each discharged patient, the key data were extracted from EMR (see eCodebook) and imported into REDCap by one of three auditors. Before importing into REDCap, the patient's unique identifier was removed while ensuring that the REDCap record ID could be mapped back to the unique patient's ID through a separately maintained file.

10% of total records were audited by all three auditors to determine inter-auditor consistency. Any discrepancies were discussed collectively, and if non-consensus or uncertainty remained, a researcher with clinical expertise was consulted.

To ensure accuracy, cases meeting any of the following criteria (related to key outcomes) underwent a second audit by an independent auditor with clinical expertise:

a. Unclear reason for discharge
b. Unclear gender identity at discharge
c. Unclear gender identity at initial appointment
d. Re-identification with birth assigned sex

Finally, where there was indication that a young person had re-identified with their birth-assigned sex, additional data pertaining to 1) timing of re-identification during their episode of care and 2) indication of regret associated with medical care were extracted from the patients EMR by an auditor and verified by a second auditor with clinical expertise. Any discrepancies were discussed, and if non-consensus or uncertainty remained, a third auditor was consulted.

We acknowledge that data collected manually remain subject to potential data extraction and data entry errors.

**Appointment data**

Appointment history was extracted directly from EMR. To determine the number of appointments a patient had received at the RCHGS, relevant departments and service providers were identified in consultation with two RCHGS clinicians and clinic administrative staff to ensure that only relevant gender-related appointments were included in the extract. Appointment data, including appointment date, attendance status, department, appointment type, and service provider, were then exported from the EMR into a CSV file using a customised purpose-built report.

The CSV file was subsequently used as a data source in Stata to analyse the data and derive 1) number of appointments from first appointment to commencement of gender affirming treatment and 2) number of appointments from first appointment to discharge.

**Regret**

First, clinical notes in the electronic medical record (EMR) were reviewed for documented expressions of regret. Second, information relating to hormone treatment was obtained from a pre-existing Hormone Audit conducted at RCHGS and supplemented with a direct EMR medication export to determine when a young person commenced hormone treatment. Finally, self-reported data were obtained from LimeSurvey questionnaires administered to young people aged 12 years and older through RCHGS and the Trans20 study.^1^ Responses relating to regret following treatment with puberty blockers, oestrogen, and testosterone were extracted using the internally developed Cessation of Hormone Treatment data extraction instrument, which captures information on hormone commencement, cessation, and any self-reported feelings of regret associated with treatment. These data were used to supplement the information obtained from the clinical records and hormone audit.

**7.0 Data Management**

Once data extraction was completed and data exports were undertaken, the Data Manager (TR) independently reviewed the compiled dataset to ensure overall data quality and consistency across variables and records. The aim was to identify and remedy issues relating to data completeness, inconsistencies, invalid or miscoded responses, and potential data entry errors not detected during the initial audit process.

**8.0 Statistical Analyses**

Statistical analyses were conducted using Stata version 18 (StataCorp).^2^ Descriptive statistics were used to summarise outcomes, including proportions for categorical data, and medians and interquartile ranges for continuous data. Calculations were performed on complete cases, i.e., missing data were excluded from calculations at the variable level.

**eCodebook**

The data recorded in REDCap (and the coding system with explanatory notes in parentheses) is provided in eTables 1-3. The data used in the current study (primary outcomes) are presented first (eTable 1), followed by data pertaining to other outcomes (eTable 2) and then supplementary data are detailed (eTable 3).

**eTable 1 – Data extracted related to primary outcomes**

| **Medical intervention** | |
| --- | --- |
| **Field** | **Code** |
| Pubertal suppression commencement whilst in the Gender Service. | 1, Yes  2, No |
| Gender affirming hormones commencement whilst in the Gender Service. | 1, Yes, estradiol  2, Yes, testosterone  3, No |
| Temporary cessation and subsequent recommencement of pubertal suppression. | 1, Yes  2, No |
| Cessation of pubertal suppression at discharge. | 1, Yes  2, No |
| Reason for cessation of pubertal suppression.** | 1, Commenced gender affirming hormones and no longer indicated  2, Adverse reaction  3, Decided to continue with endogenous puberty  4, Shift in gender identity  5, Reidentification with birth assigned sex  6, Other* |
| Date pubertal suppression commenced. | DD-MM-YYYY |
| Date pubertal suppression ceased. | DD-MM-YYYY |
| Temporary cessation and subsequent recommencement of gender affirming hormones. | 1, Yes  2, No |
| Cessation of gender affirming hormones at discharge. | 1, Yes  2, No |
| Reason for cessation of gender affirming hormones.** | 1, Reached embodiment goals  2, Adverse reaction  3, Shift in gender identity to non-binary  4, Reidentification with birth assigned sex  5, Other* |
| Date gender affirming hormone usage commenced. | DD-MM-YYYY |
| Date gender affirming hormone usage ceased. | DD-MM-YYYY |
| **Reidentification with birth-assigned sex** | |
| Gender identity at service entry. | 1, Trans masculine  (includes young people who identify as male where this differs from their birth-assigned sex)  2, Trans feminine  (includes young people who identify as female, where this differs from birth-assigned sex)  3, Non-binary  (includes young people who identify as non-binary, gender neutral, genderqueer, agender, or gender fluid, or, a combination of male/transgender male, and female/transgender female)  4, Cis-male  (includes young people who identify as male where this aligns with their birth-assigned sex)  5, Cis-female  (includes young people who identify as female where this aligns with their birth-assigned sex)  6, Young person unsure  (includes young people who are uncertain of their gender identity)  8, Unclear to auditor (this was indicated if there was ambiguous or insufficient information in the EMR for the auditor to determine gender identity).  7, Prefer not to say |
| Gender identity at service discharge. | 1, Trans masculine  (includes young people who identify as male where this differs from their birth-assigned sex)  2, Trans feminine  (includes young people who identify as female, where this differs from birth-assigned sex)  3, Non-binary  (includes young people who identify as non-binary, gender neutral, genderqueer, agender, or gender fluid, or, a combination of male/transgender male, and female/transgender female)  4, Cis-male  (includes young people who identify as male where this aligns with their birth-assigned sex)  5, Cis-female  (includes young people who identify as female where this aligns with their birth-assigned sex)  6, Young person unsure  (includes young people who are uncertain of their gender identity)  8, Unclear to auditor (this was indicated if there was ambiguous or insufficient information in the EMR for the auditor to determine gender identity).  7, Prefer not to say |
| Birth registered sex. | 1, Male  2, Female  3, Unknown |
| Reidentification with birth registered sex. | 1, Yes  2, No |
| Date of re-identification with birth registered sex. | DD-MM-YYYY |
| **Appointment information** | |
| Date of discharge*** | DD-MM-YYYY |
| Date of first appointment*** | DD-MM-YYYY |
| **Regret** | |
| Indication of regret regarding initiation of pubertal suppression. | 1, Yes  0, No |
| Further details on regret regarding initiation of pubertal suppression. | Free text. |
| Indication of regret regarding initiation of gender affirming hormones. | 1, Yes  0, No |
| Further details on regret regarding initiation of gender affirming hormones. | Free text. |
| Additional information on re-identification. | Free text. |
| *all ‘Other’ fields are followed by a ‘Specify other’ field.  ** indicates more than one option can be selected.  ***Date of first appointment and date of discharge: These dates frame one “episode of care” within the service, incorporating all the appointments and decisions made within that episode. In a minority of cases, young people had more than one episode of care with the RCHGS – defined as attending at least one appointment after a previously documented discharge. This may be for a single brief consultation, or for another longer episode of care. Some participants in the dataset may have data gathered across multiple episodes of care. | |

**eTable 2 – Data extracted related to other outcomes**

| **Discharge information** | |
| --- | --- |
| **Field** | **Code** |
| Discharge code. | 1, A - Discharged to adult or other community services  2, NM - Not seeking medical pathway  3, R - Reidentified with birth assigned sex  4, RR - Discharged and re-referred  5, NGD - Not gender diverse or cis-gender throughout  6, U - Unclear (unclear if sought ongoing gender affirming medical care) |
| Services providing care beyond discharge. ** | 1, General practitioner  2, Endocrinologist  3, Orygen Youth Health  4, Monash Gender Service  5, External psychologist  6, Other*  7, None |
| **Medical intervention** | |
| Initial type of pubertal suppression used. | 1, Leuprorelin  2, Goserelin  3, Triptorelin  4, Other* |
| Initial dosage of pubertal suppression used. | 1, 30mg Q3 monthly  2, 30mg Q4 monthly  3, 45mg Q6 monthly  4, 22.5mg Q6 monthly  5, 10.8mg Q3 monthly  6, Other* |
| Initial type of testosterone used. | 1, Testosterone topical  2, Testosterone Undecanoate  3, Testosterone Enanthate  4, Testosterone Esters |
| Initial dosage of testosterone used. | 1, 12.5mg daily  2, 25mg daily  3, 37.5mg daily  4, 50mg daily  5, 1g injection Q3 monthly  6, 250mg Q3 monthly  8, 125mg Q3 weekly  7, Other* |
| Initial type of estradiol used | 1, Oral estradiol (Progynova, Estrofem)  2, Topical estradiol (Estradot, Estraderm, Climara) |
| Initial dosage of estradiol used. | 1, 0.5mg daily  2, 1mg daily  3, 2mg daily  4, 3mg daily  5, 4mg daily  6, 12.5mcg/24hour  7, 25mcg/24hour  8, 37.5mcg/24hour  9, 50mcg/24hour  10, 75mcg/24hour  11, 100mcg/24 hour  12, Other* |
| *all ‘Other’ fields are followed by a ‘Specify other’ field.  ** indicates more than one option can be selected. | |

**eTable 3 – Supplementary data extracted**

| **Medical intervention** | |
| --- | --- |
| **Field** | **Code** |
| Pubertal suppression used. | 1, Yes  2, No |
| Initial type of pubertal suppression used. | 1, Lucrin  2, Eligard  1, Diphereline  1, Zoladex implant |
| Reason for temporary cessation of pubertal suppression. | Free text |
| Further description of reason for cessation of pubertal suppression. | Free text |
| Final type of pubertal suppression used. | 1, Leuprorelin  2, Goserelin  3, Triptorelin  4, Other* |
| Final dosage of pubertal suppression used. | 1, 30mg Q3 monthly  2, 30mg Q4 monthly  3, 45mg Q6 monthly  4, 22.5mg Q6 monthly  5, 10.8mg Q3 monthly  6, Other* |
| Final type of pubertal suppression used. | 1, Lucrin  2, Eligard  1, Diphereline  1, Zoladex implant |
| Gender affirming hormone used. | 1, Yes, estradiol  2, Yes, testosterone  3, No |
| Reason for temporary cessation of gender affirming hormones. | Free text |
| Further description of reason for cessation of gender affirming hormones. | Free text |
| Final type of estradiol used. | 1, Oral estradiol (Progynova, Estrofem)  2, Topical estradiol (Estradot, Estraderm, Climara) |
| Final dosage of estradiol used. | 1, 0.5mg daily  2, 1mg daily  3, 2mg daily  4, 3mg daily  5, 4mg daily  6, 12.5mcg/24hour  7, 25mcg/24hour  8, 37.5mcg/24hour  9, 50mcg/24hour  10, 75mcg/24hour  11, 100mcg/24 hour  12, Other* |
| Final type of testosterone used. | 1, Testosterone topical  2, Testosterone Undecanoate  3, Testosterone Enanthate  4, Testosterone Esters |
| Final dosage of testosterone used. | 1, 12.5mg daily  2, 25mg daily  3, 37.5mg daily  4, 50mg daily  5, 1g injection Q3 monthly  6, 250mg Q3 monthly  8, 125mg Q3 weekly  7, Other* |
| Use of progestogens as part of feminising hormone treatment. | 1, Yes  2, No |
| Progestogen type used. | Free text. |
| Progestogen dose used. | Free text. |
| Progestogen date commenced. | DD-MM-YYYY |
| Progestogen date ceased. | DD-MM-YYYY |
| Use of antiandrogens. | 1, Yes  2, No |
| Commencement of antiandrogens whilst in the RCH Gender Service. | 1, Yes  2, No |
| Initial type of antiandrogens used. | 1, Cyproterone acetate PO  2, Spironolactone |
| Initial dose of antiandrogens used. | 1, 12.5mg twice per week  2, 12.5mg daily  3, 25mg daily  4, 100mg PO daily  5, 200mg PO daily  6, Other* |
| Date antiandrogens commenced | DD-MM-YYYY |
| Temporary cessation and subsequent recommencement of antiandrogen usage. | 1, Yes  2, No |
| Reason for temporary cessation of antiandrogen usage. | Free text |
| Cessation of antiandrogen usage at discharge. | 1, Yes  2, No |
| Date antiandrogen usage ceased | DD-MM-YYYY |
| Reason for antiandrogen usage cessation.** | 1, Reached embodiment goals  2, Adverse reaction  3, Shift in gender identity to non-binary  4, Reidentification with birth assigned sex  5, Other* (please type reason) |
| Further description of reason for antiandrogen usage cessation. | Free text |
| Final type of antiandrogen used. | 1, Cyproterone acetate PO  2, Spironolactone |
| Final dose of antiandrogen used. | 1, 12.5mg twice per week  2, 12.5mg daily  3, 25mg daily  4, 100mg PO daily  5, 200mg PO daily  6, Other* |
| Gender affirming surgery*** undertaken at external service provider prior to discharge.** | 1, Top surgery  2, Vaginoplasty  3, Orchidectomy without vaginoplasty  4, Other* |
| Date of top surgery | DD-MM-YYYY |
| Date of vaginoplasty | DD-MM-YYYY |
| Date of orchidectomy without vaginoplasty | DD-MM-YYYY |
| Date of Other* surgery | DD-MM-YYYY |
| **Appointment Information** | |
| Single-session nurse assessment clinic appointment attended. | 1, Yes  2, No |
| Multidisciplinary clinic appointment attended. | 1, Yes  2, No |
| Paediatrician or endocrinologist for gender-related appointment attended. | 1, Yes  2, No |
| Mental health clinician for gender-related appointment attended. | 1, Yes  2, No |

*all ‘Other’ fields are followed by a ‘Specify other’ field.

** Indicates more than one option can be selected.

*** Gender affirming surgery is not provided at RCHGS

1. Prior to the establishment of the RCH Gender Service, children and adolescents could be seen by individual clinicians at the RCH for gender-related matters.
